# Assess Alzheimer’s Disease via Plasma Extracellular Vesicle-derived mRNA

**DOI:** 10.1101/2023.12.26.23299985

**Authors:** Le Hoang Phu Pham, Ching-Fang Chang, Katherine Tuchez, Yuchao Chen

## Abstract

Alzheimer’s disease (AD), the most prevalent neurodegenerative disorder globally, has emerged as a significant health concern, particularly due to the increasing aging population. Recently, it has been revealed that extracellular vesicles (EVs) originating from neurons play a critical role in AD pathogenesis and progression. These neuronal EVs can cross the blood-brain barrier and enter peripheral circulation, offering a less invasive means for assessing blood-based AD biomarkers. In this study, we analyzed plasma EV-derived messenger RNA (mRNA) from 82 subjects, including individuals with AD, mild cognitive impairment (MCI), and healthy controls, using next-generation sequencing (NGS) to profile their gene expression for functional enrichment and pathway analysis. Based on the differentially expressed genes identified in both MCI and AD groups, we established a diagnostic model by implementing a machine learning classifier. The refined model demonstrated an average diagnostic accuracy over 98% and showed a strong correlation with different AD stages, suggesting the potential of plasma EV-derived mRNA as a promising non-invasive biomarker for early detection and ongoing monitoring of AD.

## Introduction

The pursuit of early AD diagnosis is essential to combat cognitive decline with timely intervention and symptomatic treatment before irreversible brain damage.^1–3^ Integrative diagnostic approaches now employ a combination of cognitive and functional assessment, biomarker identification, and neuroimaging to uncover the earliest signs of AD.^4^ However, the high costs of positron emission tomography (PET) imaging restrict its widespread use in clinical settings, especially in under-resourced areas. While cerebrospinal fluid (CSF) is considered the optimal source for AD biomarkers due to its direct interaction with the brain, the invasiveness and potential complications of lumbar puncture, required for CSF collection, limit its frequent use.^5^ Researchers have been exploring the potential of identifying non-invasive AD biomarkers in alternative body fluids, particularly blood.

Neuronal extracellular vesicles (EVs) have recently been recognized as significant contributors to AD pathology.^6^ These membrane-bound nano vesicles, released by neurons, encapsulate a range of cellular components including nucleic acids, proteins, and lipids, that reflect their originating cells.^7^ In AD, neuronal EVs are known to carry hallmark biomolecules associated with the disease, indicating their potential role in spreading pathology within the brain.^8^ Furthermore, these vesicles are implicated in the modulation of neuroinflammation, a key component of AD’s pathological progression.^9^ Beyond their role in disease mechanisms, neuronal EVs are gaining attention as potential diagnostic tools.^10, 11^ Their stability and presence in various biofluids, such as blood, offer a minimally invasive window into the neuronal state and AD-related changes. Therefore, analyzing the content of neuronal EVs could provide insights into the early stages and progression of AD, highlighting their value in diagnosis.

Recent studies have leveraged RNA sequencing or microarray technology to explore the contents of EVs, leading to the identification of potential biomarkers for early diagnosis of AD. Extensive studies have focused on EV-derived micro-RNA (miRNA) obtained from various sources such as brain tissues, cerebrospinal fluid (CSF), and blood. One study by Cheng *et al.* revealed an upregulation of disease-associated miRNA within EVs derived from AD-affected brain tissues.^12^ This finding suggests neuronal EVs could serve as viable indicators for early-stage AD, facilitating a non-invasive diagnostic approach. Furthermore, other research has identified distinct miRNA expression patterns in plasma-derived EVs.^13–17^ Some patterns are not only unique to AD but also capable of differentiating it from other forms of dementia, thereby enhancing the specificity of diagnostic tools. These findings emphasize the importance of EV-derived small RNA in the pathogenesis and potential diagnosis of AD.

In addition to micro-RNA (miRNA), messenger RNA (mRNA) in neuronal EVs may also play a crucial role in AD pathology. mRNA, carrying genetic information for protein synthesis, is found within EVs and can be transferred to recipient cells, where it is translated into proteins that alter cellular behavior.^18^ The secretion of EVs occurs not only in various normal physiological states but also in pathological conditions. Of particular interest is the recent focus on EV-carried mRNAs in cancer research, where they have been found to play significant roles in tumor progression and the modification of the tumor microenvironment.^19^ The importance of mRNA in AD is similarly noteworthy as it offers direct insights into gene expression patterns. However, research specifically targeting EV-derived mRNA in AD is still relatively sparse. Notably, Luo *et al*. conducted a study profiling long RNA within EVs derived from AD brain tissues.^20^ Their findings revealed a substantial number of differentially expressed genes (DEGs), some of which are linked with transcriptional changes observed in AD. These DEGs, upon analysis, could shed light on alterations in protein translation processes, which are indicative of the onset and progression of the disease. Given these insights, profiling EV-derived mRNA may pave the way for novel diagnostic and therapeutic approach in the management of AD.

This study aims to further our understanding of AD and improve its early diagnostic capabilities through the comprehensive profiling of mRNA from plasma EVs. Neuronal EVs, which can cross the blood-brain barrier while maintaining cargo stability, offer a non-invasive avenue to investigate the molecular signatures associated with this disease. Leveraging NGS, we analyzed mRNA libraries derived from EVs in 82 plasma samples, including 44 individuals at various AD stages, 13 with MCI who later progressed to AD, and 25 healthy controls. Our analysis revealed differential gene expression patterns, enabling in-depth exploration of DEGs associated with AD. Gene Ontology (GO) and Kyoto Encyclopedia of Genes and Genomes (KEGG) pathway analyses provide valuable insights into the neurodegenerative and metabolic pathways influenced by these genes. Additionally, by comparing gene expression profiles from both AD and MCI cases with those from healthy controls, we identified a collection of gene biomarker candidates. These candidates underwent rigorous functional and correlation analyses, ultimately resulting in the establishment of a classification model based on a support vector machine (SVM), showing high accuracy and strong correlation with the clinical dementia rating (CDR) scale. In summary, our study developed a biomarker panel with an AD classification model based on plasma EV-derived mRNA, shedding light on AD pathophysiology and enhancing its diagnostic potential.

## Results

### Integrated transcriptome and pathway analysis

As shown in Figure 1A, we isolated and purified small EVs from 0.4 mL of plasma specimens, collected from a cohort of 82 individuals. The specimen information is summarized in **Table S1**. Following this, mRNA was extracted from these EV samples for sequencing library preparation. Subsequent RNA sequencing and gene alignment led to the identification of 19,495 genes. Categorization by biotype revealed the presence of 15,664 protein-coding genes (80.34%), 2,391 long non-coding RNAs (12.26%), and 1,440 genes of other biotypes (7.38%) within the identified dataset. This dataset, notably enriched with protein-coding genes, forms the foundation for the analysis conducted in our study. The volcano plots of the differential expression analysis within the MCI group (Figure 1B) revealed 1,097 significantly expressed protein-coding genes when compared to the Control group, including 897 upregulated and 200 downregulated genes. In the AD group (Figure 1C), 1,701 protein-coding genes were found to be significantly differentially expressed in comparison to the Control group, with 1,421 upregulated and 280 downregulated. We analyzed differentially expressed genes (DEGs) in each group to assess their involvement in biological pathways, utilizing g:Profiler against the KEGG database. The analysis indicated significant enrichment in the AD pathway. Specifically, in the MCI group, we observed an enrichment with a p-adjusted value of 1.36e-7, involving 27 AD-related genes, which have been identified in g:Profiler. In the AD group, the enrichment was more pronounced, with a p-adjusted value of 1.25e-13 and 44 AD-related genes. These genes are labeled in Figures 1B and **1C**. Furthermore, we visualized the expression levels of these AD-related genes through heatmaps in Figures 1D and **1E**. The heatmaps display distinct expression patterns in comparison to Control samples for both MCI and AD cases, offering insightful visual representation of the data.

**Figure 1.**
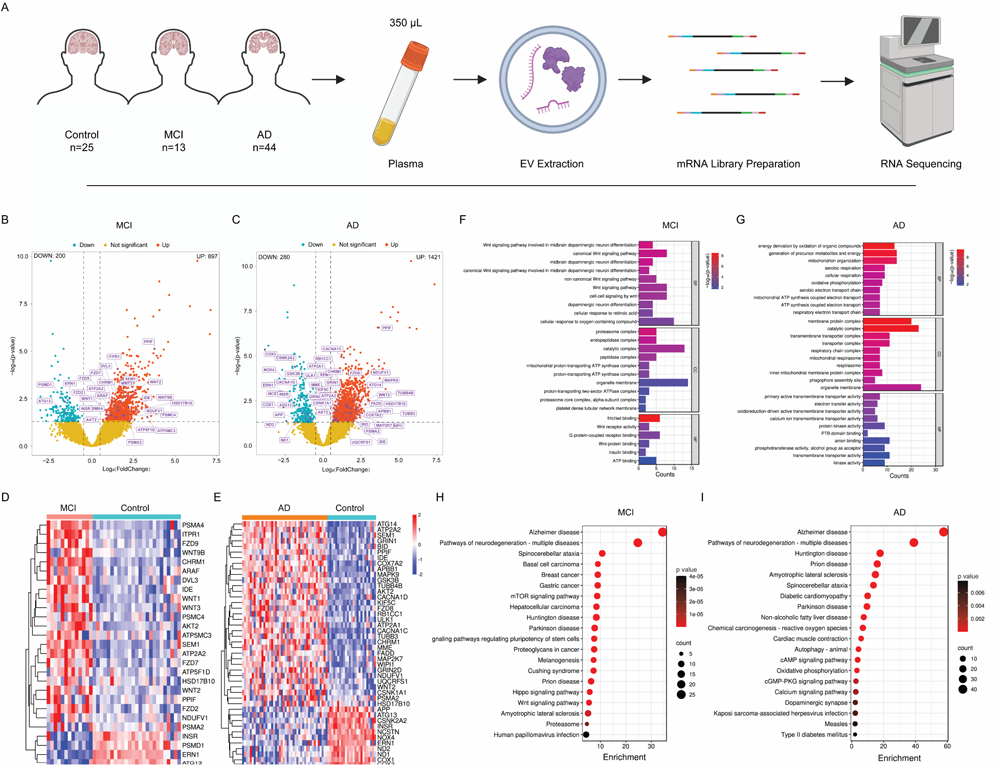
Analysis of mRNA from plasma-derived EVs highlighting key AD-related genes in MCI and AD groups. (**A**) Illustration of sample-preparation workflow. (**B-C**) Volcano plots showing differentially expressed protein-coding genes in MCI and AD groups compared to the Control group (p-adjusted < 0.05 and log2 fold change > |0.5|). The labeled genes are AD-related. (**D-E**) Heatmaps display expression patterns of the AD-related DEGs in MCI and AD groups compared to the Control group. Gene clustering in the heatmaps was based on hierarchical clustering using the Euclidean distance metric. (**F-G**) GO annotations for the AD-related DEGs in MCI and AD groups. (**H-I**) KEGG pathway enrichment analysis for AD-related DEGs in MCI and AD groups.

We conducted a comparison analysis of GO annotations for AD-related DEGs in the MCI (Figure 1F) and AD (Figure 1G) groups, focusing on the top 10 most significant annotations in each category. In the Biological Processes (BP) category, DEGs in MCI group predominantly engage with the Wnt signaling pathway, with 8 genes in both canonical and general Wnt pathways, and 10 genes in cellular response to oxygen-containing compounds. On the other hand, in the AD group, DEGs are mainly involved in energy metabolism pathways, such as oxidation of organic compounds (13 genes) and generation of metabolites and energy (14 genes). In the Cellular Components (CC) domain, MCI DEGs are primarily associated with protein complexes, including the catalytic complex (13 genes) and organelle membrane (14 genes). For AD group, the emphasis shifted to cellular structures like organelle membranes (24 genes) and catalytic complexes (23 genes). Within the Molecular Functions (MF) category, MCI genes were involved in binding activities, including frizzled binding (6 genes) and G protein-coupled receptor binding (6 genes). In contrast, AD genes focused on enzymatic functions, such as transmembrane transporter (11 genes), kinase (9 genes), and protein kinase activities (9 genes). This transition reflects the shift in molecular characteristics and functional emphasis from MCI to AD.

We subsequently conducted a KEGG pathway analysis of AD-related DEGs in both groups, revealing similarities and distinctions between the two conditions. The top 20 significant pathways for MCI (Figure 1H) and AD (Figure 1I) provide a detailed insight into the genetic landscape and underlying molecular mechanisms of both states. A key observation was the overlap in several pathways, including Alzheimer’s disease, neurodegeneration-related pathways, and other neurologically-linked diseases such as Spinocerebellar ataxia and Parkinson’s disease, common to both MCI and AD. Notably, the enrichment data indicated a stronger association with the AD group, especially in the Alzheimer’s disease pathway, where enrichment scores were significantly higher at 57.69 for AD group compared to 34.50 for MCI group. The difference suggests a progressive genetic intensification from MCI to AD state. While these shared genetic pathways revealed their interconnected nature, distinct pathways unique to each condition were also identified. MCI showed a tendency towards pathways typically linked to cellular processes and carcinomas, such as the mTOR signaling pathway, pluripotency of stem cells, and Melanogenesis. In contrast, AD was more aligned with pathways influencing metabolic and systemic processes, including Diabetic cardiomyopathy, Oxidative phosphorylation, and the Calcium signaling pathway. A further analysis of gene counts within these pathways revealed a consistent increase in gene numbers in AD group. For example, in the Huntington disease pathway shared by both conditions, AD group had a higher gene count with 22 genes compared to 12 genes in MCI group. This pattern indicates a potential accumulation of genes in AD, thereby amplifying its genetic complexity relative to MCI.

### Identification of potential AD biomarkers

In the study to identify biomarkers for AD, we focused on a subset of DEGs shared by MCI and AD groups (Figure 2A), which made up 22.5% of the total AD-related gene pool. This indicates a notable genetic overlap between MCI and AD, suggesting these genes are potentially critical in AD progression. The expression patterns of these genes, analyzed through variance-stabilizing transformation (VST), showed distinct trends: 10 genes were upregulated and 3 were downregulated in MCI and AD stages compared to the Control group. These trends were quantified statistically as shown in Figure 2B. Generally, the upregulated genes showed higher median VST values in diseased states, while downregulated genes had lower median VST values, indicating a reduction in expression as the disease advances. An exception was PSMA2, which did not exhibit a clear up or downregulation trend when MCI and AD were compared with the Control group. The statistical significance of these differential expressions was marked with asterisks above the boxplots (Figure 2B).

**Figure 2.**
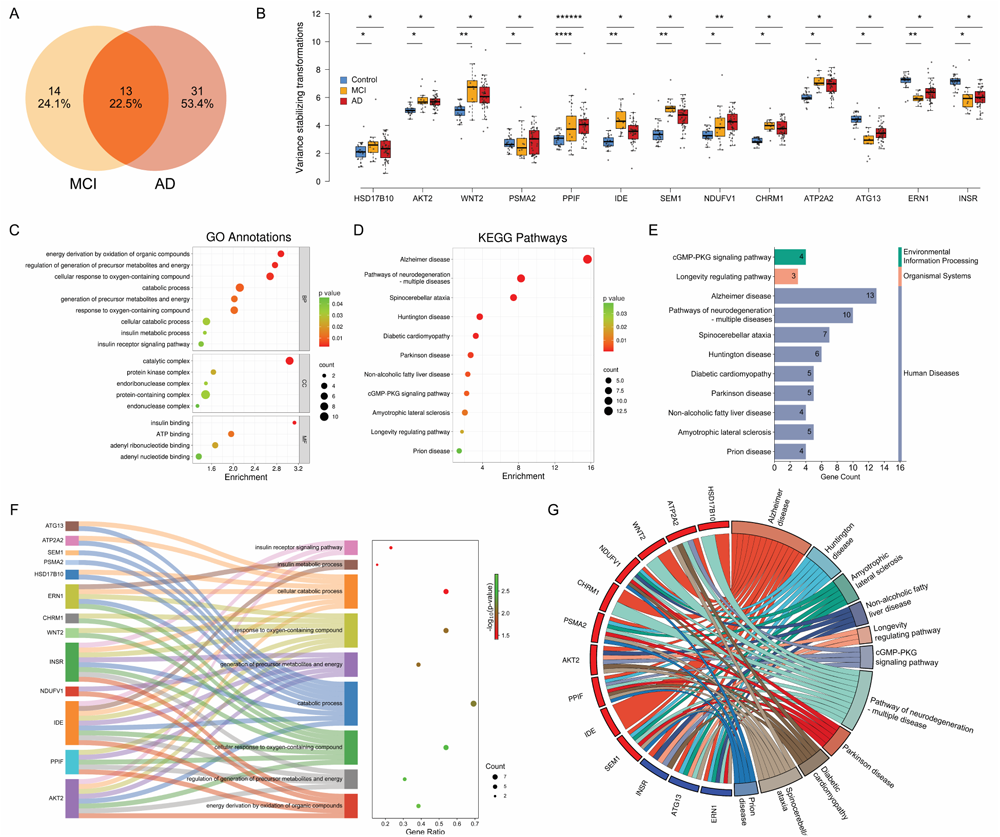
Integrative analysis of 13 shared DEGs. (**A**) Venn diagram showing the overlap of differentially expressed AD-related genes between the MCI and AD groups. (**B**) Box plot showing variations in gene expression across the 13 shared AD genes through different disease stages: Control, MCI, and AD. (*p < 0.05, **p < 0.005, ****p < 0.00005, ******p < 0.0000005) (**C**) Enrichment bubble plot illustrating the GO annotations for the 13 genes. (**D**) Enrichment bubble plot illustrating the KEGG pathways enriched by the 13 genes. (**E**) Summary of pathway enrichment listing the number of genes involved in each pathway. (**F**) Sankey diagram mapping the 13 genes to their associated biological processes. (**G**) Chord plot showing the relationships between the 13 genes and their linked KEGG pathways.

### Functional enrichment and pathway analysis of common genes

We further conducted GO annotations for the 13 AD-related DEGs shared between MCI and AD groups (Figure 2C). The analysis ranked these annotations based on their enrichment scores relative to the background dataset. The significance of these findings was further confirmed by corresponding p-adjusted values. Within the Biological Process (BP) category, there was a significant enrichment in processes linked to cellular metabolism, including energy derivation through oxidation of organic compounds and regulation of precursor metabolites and energy generation, engaging 5 and 4 genes respectively. Notably, pathways related to cellular response to oxygen-containing compounds and response to these compounds involved 7 genes each, suggesting a potential connection to oxidative stress responses in neurodegenerative pathogenesis. In the Cellular Component (CC) domain, the protein-containing complex was prominent, with 11 genes involved, suggesting a major role of these molecules in AD-related cellular changes. The catalytic complex, enriched with 8 genes, emphasized the importance of enzymatic activities in the AD cellular environment. Additionally, the protein kinase complex and endoribonuclease complex were also highlighted, indicating a complex interaction of various protein complexes in the disease development. For Molecular Function (MF) annotations, insulin binding emerged as highly significant, involving 2 genes, indicating potential disruptions in insulin signaling pathways in AD. This is accompanied by nucleotide-binding functions such as ATP binding, adenyl ribonucleotide binding, and adenyl nucleotide binding, each implicating 4 genes. These findings suggest alterations in energy dynamics and signaling processes in AD.

Figure 2F shows a deeper insight into the distribution and roles of the 13 AD-related DEGs in Biological Process (BP), particularly in the pathways critical to cellular metabolism and the body’s response to oxidative stress. The Sankey diagram mapped the connection of these genes to specific biological processes, emphasizing both their gene ratio and p-adjusted significance. The analysis revealed a significant enrichment in the pathway for energy derivation through the oxidation of organic compounds, involving 5 genes: AKT2, PPIF, IDE, NDUFV1, and INSR. This pathway shows a notable p-adjusted value of 0.0013, confirming its significance. Additionally, the regulation of the generation of precursor metabolites and energy, which includes 4 genes (AKT2, PPIF, IDE, and INSR) with a p-adjusted value of 0.0017, further highlights the crucial role of these genes in key metabolic pathways that are often disrupted in AD.^21, 22^ Moreover, the pathways related to cellular responses to oxygen-containing compounds are notable as well, each involving 7 genes - AKT2, WNT2, PPIF, IDE, CHRM1, INSR, and ERN1. With a gene ratio of 0.538 and identical p-adjusted values of approximately 0.0021, these pathways emphasize the collective role of these genes in the oxidative stress response, which is crucial in the pathogenesis of neurodegenerative diseases like AD.^23–25^

The KEGG pathway analysis of 13 AD-related DEGs is shown in Figure 2D, ranked by their enrichment scores. Notably, the Alzheimer’s disease pathway, with a significant enrichment score of 15.625 and a remarkably low p-adjusted value of 2.37e-16, emerged at the top of the list. This pathway, involving all 13 genes, emphasized the common genetic basis from MCI to AD. The detailed visualization of this pathway is illustrated in **Figure S1**, showing the involvement of these genes in a range of AD-associated biological processes. The ’Pathways of Neurodegeneration - Multiple Diseases’ follows with an enrichment score of 8.265, involving 10 genes, emphasizing their broader impact on various neurodegenerative diseases beyond Alzheimer’s. The Spinocerebellar ataxia and Huntington’s disease pathways also showed notable enrichment scores of 7.437 and 3.693, respectively, indicating genetic overlaps with these specific neurodegenerative disorders. Further, pathways like Diabetic Cardiomyopathy, Parkinson’s Disease, and Non-alcoholic Fatty Liver Disease, with enrichment scores of 3.257, 2.688, and 2.371, respectively, suggested the AD-related genes’ relevance to metabolic and systemic diseases, linking Alzheimer’s pathology with wider metabolic issues. Additionally, signaling pathways like the cGMP-PKG signaling pathway, key for vascular and neuronal functions, and the ’Longevity Regulating Pathway’, associated with aging, are also highlighted in the analysis with enrichment scores of 2.244 and 1.734. Even lesser-scored pathways, such as Amyotrophic Lateral Sclerosis and Prion Disease, demonstrated the implicated genes’ involvement in various disease mechanisms. Categorization of the pathways in Figure 2E indicated the significance of these genes in human diseases, particularly neurodegenerative disorders. The distribution of genes across these pathways, as reflected in the gene counts, aligns with the level of enrichment, painting a comprehensive picture of these genes’ roles in disease-related biological processes.

Figure 2G presents a chord diagram that maps the complex relationships between genes and pathways, providing a visualization of their regulatory impact. Notably, gene ATP2A2, interacting with various pathways including Diabetic cardiomyopathy and Spinocerebellar ataxia, indicated the possible wide-ranging effects of certain genes on both neurodegenerative and metabolic diseases. This suggests that the dysregulations observed in Alzheimer’s could have extensive systemic effects, potentially influencing or being influenced by peripheral tissue dysfunction.^26, 27^ NDUFV1, PSMA2, and SEM1 showed broad engagement with pathways such as Huntington disease, Amyotrophic lateral sclerosis, and Prion disease. Their upregulated expression in these diverse neurodegenerative conditions might suggest a common pathological mechanism or a unified response to neurodegenerative stress.^28^ AKT2 and PPIF also exhibited wide-ranging associations with multiple pathways. AKT2’s involvement in these pathways reflected its dual role in the central nervous system’s pathology and peripheral metabolic disturbances related to AD.^26^ PPIF’s widespread links suggested its role in mitochondrial dysfunction, a hallmark of neurodegenerative diseases.^29^ The genes INSR, ATG13, and ERN1 display a diverse array of connections as well. INSR’s links to these pathways suggested that insulin resistance, a feature often seen in metabolic syndrome, may also play a role in neuropathology of AD.^30, 31^ On the other hand, the ties of ATG13 and ERN1 to longevity and mitochondrial pathways emphasized the significance of cellular aging and energy regulation in the progression of AD.^32, 33^ These correlations are enlightening as they reveal the intersection of metabolic regulation, aging, and neurodegeneration, suggesting that genes involved in AD could be crucial in these broader biological processes.

### Biomarker selection through correlation analysis and clustering

We conducted a refinement of the 13 AD-related DEGs through biweight midcorrelation analysis to characterize the similarity in expression among the biomarker candidates. This approach identified gene pairs with a high degree of correlation, specifically those with a correlation coefficient above |0.65|. For instance, ATP2A2 and SEM1, as shown in Figure 3A, were closely aligned, suggesting potential exclusion to maintain dataset integrity and avoid redundancy. Additionally, the dataset was assessed through a comprehensive clustering analysis using Euclidean distance, resulting in a heatmap shown in Figure 3B. This heatmap clustered genes based on their expression patterns across all diagnostic categories: AD, MCI, and Control. A dendrogram on the heatmap shows the hierarchical relationships between genes, aiding in identifying closely related pairs. To minimize dataset redundancy, gene pairs that clustered closely, such as ATG13 with INSR, PSMA2 with HSD17B10, NDUFV1 with PPIF, WNT2 with CHRM1, and SEM1 with ATP2A2, were marked for potential exclusion. The goal was to select one gene from each pair for the final biomarker panel, ensuring a unique and non-overlapping set for further validation.

**Figure 3.**
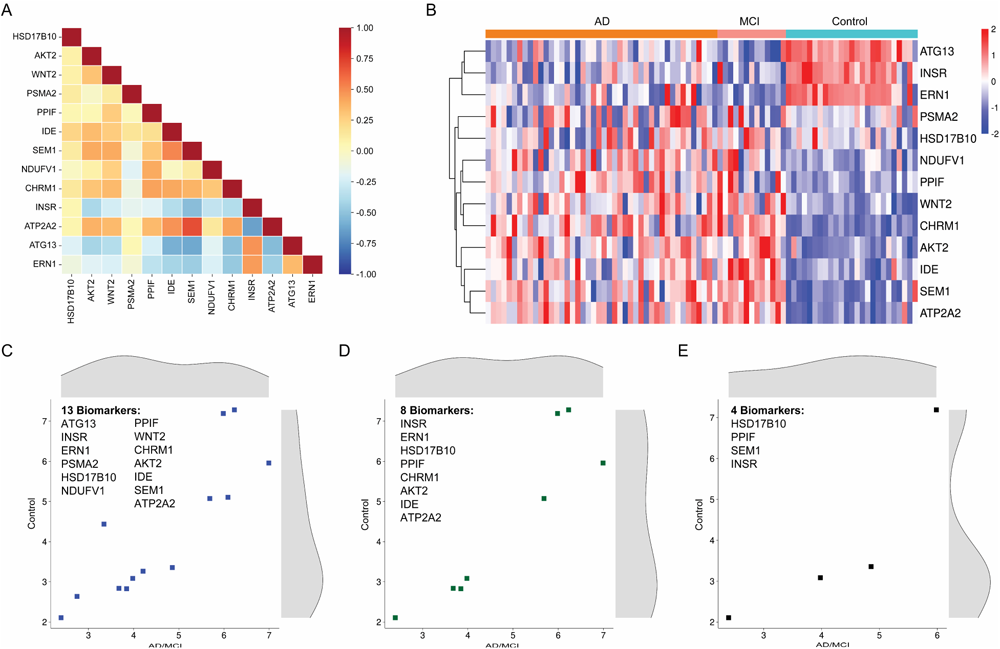
Biomarker selection through correlation analysis and clustering. (**A**) Biweight correlation matrix showing genes with strong correlations. Color intensity represents the correlation strength with darker hues indicating stronger correlations. (**B**) Heatmap with a dendrogram constructed using Euclidean distance, showing gene expression similarities among the 13 AD genes. (**C-E**) Scatter plots showing the median expression distributions of the Control and AD/MCI groups across three selected gene sets.

Following the analysis, we derived three potential biomarker panels for further examination. As illustrated in Figures 3C-E, each panel is characterized with a scatter plot showing the median expression values for the two diagnostic categories, the Control group and a combined group of MCI and AD. Panel 1 (Figure 3C), the unrefined group, includes all the 13 genes. Panel 2 (Figure 3D), consisting of 8 genes, was created by removing one gene from each redundant pair identified in the clustering assessment. An alternative to panel 2 (**Figure S2A**), also containing 8 genes, was formed by choosing the alternate gene from each pair that was omitted in panel 2, serving as a comparison. The last panel (Figure 3E) was compiled by selecting one gene from the dendrogram branch of downregulated genes and three from the initial splits of upregulated genes, ensuring diversity of gene expression patterns in the biomarker panel. The scatter plots showed that the selected biomarker panels maintained a similar range of gene expression, allowing comprehensive coverage of potential biomarkers while minimizing redundancy, thereby enhancing their overall applicability in the subsequent validation stage.

### Development and characterization of AD prediction model

For each biomarker panel, we developed an AD prediction model by training a linear Support Vector Machine (SVM) classifier with the dataset. A nested cross-validation (5-fold inner and outer cross-validation) was first employed to determine the optimal hyperparameters for the SVM model. Subsequently, both the AD/MCI and control groups were randomly divided into a training set (80%) and a testing set (20%) each time for 500 trials of model training and testing, in order to characterize model performance in terms of Receiver Operating Characteristic (ROC) Area Under Curve (AUC) value. Figure 4A-C illustrate the mean ROC curves for each biomarker panel.

**Figure 4.**
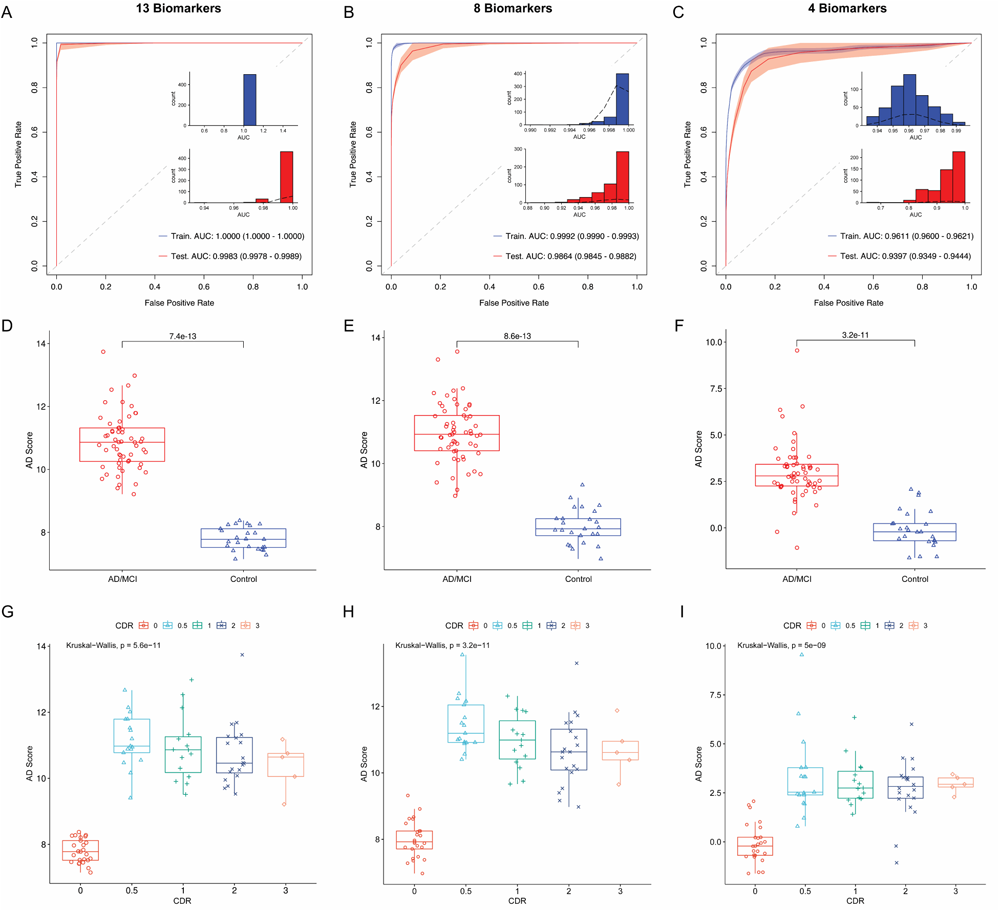
Model characterization for AD evaluation. (**A-C**) Training and testing of an SVM classification model for each biomarker panel with the performance characterized through ROC curves, highlighting the AUC and 95% confidence interval. Inset figures present the distribution of AUC values over 500 iterations, illustrating the model’s performance consistency. (**D-F**) Comparison of AD scores for each sample as generated by the prediction models, differentiating between AD/MCI and control groups. The significance of differences was characterized using the Wilcoxon test. (**G-I**) Evaluation of correlation between AD scores and CDR values. Statistical differences among groups were assessed using the Kruskal-Wallis test.

The shading of the curves indicates the standard deviations across the 500 training and testing iterations, reflecting the consistency of the models’ performance. Additionally, the distribution of AUC values (inset of Figure 4A-C) provides insights into the variation of the models’ prediction outcomes over these iterations.

The first biomarker panel (Figure 4A), incorporating all the 13 genes, displayed outstanding training performance with an AUC of 1.00. This high level of accuracy continued in the testing phase, where it obtained a mean AUC of 0.9983, with a tight 95% confidence interval (CI) between 0.9978 and 0.9989, indicating strong predictive reliability. The second biomarker panel (Figure 4B), a refined cohort consisting of 8 biomarkers, recorded a mean training AUC of 0.9992, slightly lower than that of the 13-gene set. In the testing, it reached a mean AUC of 0.9864, with a 95% CI stretching from 0.9845 to 0.9882, suggesting a slight reduction in classification capability compared to the first panel, but it still retained high diagnostic precision. An alternative version of the second panel (**Figure S2B**), also with 8 biomarkers, had a comparable but lower AUC for both training and testing to the original second biomarker panel. The final biomarker panel (Figure 4C), which included only 4 genes, exhibited a training AUC of 0.9611. Although lower compared to the other panels, it still maintained a strong capacity to differentiate between conditions. For this biomarker panel, the testing phase yielded a mean AUC of 0.9397, accompanied by a wider 95% CI ranging from 0.9349 to 0.9444, reflecting greater variability in the model’s predictive accuracy.

The AD classifier for each marker panel was developed by averaging the weights and bias terms from the linear SVM model across 500 iterations, as detailed in Supplementary Information. This process yielded a final model that generated an AD score for each sample. These scores were then compared between two groups (AD/MCI and Control), as illustrated in Figures 4D-F and **Figure S2C**. Generally, it was observed that as the number of genes in the biomarker panels reduced from 13 to 4, the overlap in the interquartile ranges of the AD/MCI and Control groups became more significant. This trend was confirmed by an increase in the p-value from the Wilcoxon test, indicating less distinction between the groups. However, the model retained significant discriminative ability even with only 4 gene markers (Figure 4F), as evidenced by a statistically significant p-value of 3.2e-11.

The prediction model not only demonstrated the ability to discriminate between the Control and AD/MCI groups but also exhibited a strong correlation with the clinical dementia rating (CDR). We subdivided the samples into various groups based on their CDR values to compare their AD scores, as shown in Figures 4G-I and **Figure S2D**. As anticipated, there was a significant increase in the median AD scores, progressing from the control group (CDR 0) to the AD/MCI group (CDR 0.5-3). More importantly, the median scores showed a gradual decrease as the CDR value increased from 0.5 to 3. This trend was consistently observed across the three biomarker panels, indicating the model’s ability to differentiate among AD stages.

## Discussion

In this study, mRNA extracted from plasma EVs of 82 subjects was analyzed, including patients with AD, individuals with MCI who later progressed to AD, and healthy controls (Figure 1A). We employed NGS to profile their gene expression for functional enrichment and pathway analysis. The findings, which revealed an increase in the number of differentially expressed genes (DEGs) from MCI to AD (Figure 1B **and** **1C**), suggest a genetic progression of the disease. The KEGG analysis showed an enrichment in the Alzheimer’s pathway (Figure 1H **and** **1I**), confirming the clinical significance of these genes as potential AD biomarkers. Furthermore, the pathways common to both MCI and AD groups, such as those related to neurodegeneration, highlight a genetic continuum, indicating that MCI is an early stage of AD.^1, 2^ This could be crucial for early diagnosis and ongoing monitoring of AD.

The GO analysis revealed distinct biological activities and molecular functions specific to MCI and AD groups (Figure 1F **and** **1G**). The prevalence of the Wnt signaling pathway in the MCI group, and the emphasis on metabolic pathways in the AD group, point to unique disease mechanisms, that may provide insights for developing targeted therapeutic strategies. The results indicate a progression in cellular complexity from MCI to AD, suggesting possible changes in cellular dynamics that could influence disease progression and the efficacy of treatments. By exploring differences in pathway activation, like the association of MCI with cellular processes and AD’s link to metabolic disturbances, these findings are consistent with growing evidence that positions AD not just as a cognitive disorder but also as a systemic disease.^34–36^

The identification of shared AD-related DEGs as potential biomarkers provides new opportunities for early detection and monitoring of disease progression. The 22.5% gene overlap indicates a substantial genetic intersection between MCI and AD groups, highlighting the potential for these biomarkers in understanding and tracking the disease. The functional enrichment and pathway analyses (Figure 2) present a biological narrative where cellular metabolism and response to oxidative stress emerge as central themes. These enriched pathways emphasize the potential disruption in metabolic processes and the crucial role of oxidative stress in neurodegenerative diseases.^37, 38^ The notable involvement of protein complexes in our study points to changes in cellular structure, while the significant presence of insulin and nucleotide binding indicates disruptions in signaling pathways that may contribute to the pathology of AD.^39, 40^ The Sankey diagram in Figure 2F illustrates how these shared genes are interconnected within a complex biological network. Key pathways, such as the energy derivation from the oxidation of organic compounds, highlight the metabolic disturbances characteristic of AD. This supports existing literature that identifies metabolic dysfunction as a crucial factor in AD’s pathogenesis. Furthermore, the genes’ collective role in oxidative stress responses sheds light on their potential impact in either mitigating or exacerbating cellular damage in AD.^21, 41^

The KEGG pathway analysis in Figures 2D and **2E** adds a comparative and quantitative perspective, placing the shared AD-related genes against the broader background of neurodegenerative and metabolic diseases. This analysis not only reaffirms the genetic basis of AD, but also hints at a genetic tendency towards systemic diseases in individuals with AD, suggesting that AD’s impact might extend beyond cognitive decline.^42^ The chord diagram in Figure 2G provides a graphic illustration of gene-pathway interactions, highlighting the diverse roles of genes like ATP2A2 and their potential to blur the boundary between neurodegenerative and metabolic disorders. This interconnectedness implies that the pathophysiological processes of AD could be closely intertwined with overall systemic health.

The extensive involvement of genes such as NDUFV1, PSMA2, and SEM1 in various neurodegenerative pathways indicates a common pathogenic mechanism. At the same time, the varied associations of AKT2 and PPIF point to their roles in both central and peripheral disease processes. Exploring the functions of INSR, ATG13, and ERN1 reveals a meeting point of metabolic regulation, aging, and neurodegeneration. This paints a picture of AD as a complex condition where metabolic imbalances and cellular aging may merge, driving the progression of the disease.

The ROC analysis of the AD prediction model exhibits outstanding performance in both training and testing, revealing the biomarkers’ effectiveness in differentiating AD patients from healthy individuals. The first biomarker panel (Figure 3C), which includes all the 13 shared genes, shows the highest accuracy (AUC>0.99). Nevertheless, the considerable similarity in gene expression raises concerns about overfitting, a scenario where the model exhibits excellent performance during the training phase but may underperform in practical, real-world applications. Upon eliminating gene redundancy, the second biomarker panel with 8 genes (Figure 3D) continues to maintain an impressive AUC greater than 0.98, making it a more credible selection for practical application. While the third biomarker panel, comprising only 4 genes, presents an appealing simplicity, it falls short in providing a broad genetic scope, resulting in reduced accuracy and greater variability during testing. Therefore, the second panel, which includes the genes INSR, ERN1, HSD17B10, PPIF, CHRM1, AKT2, IDE, and ATP2A2, stands out as the suitable choice. It achieves a more effective balance between genetic comprehensiveness and clarity of expression, thereby optimizing predictive accuracy of the model while minimizing the risk of overfitting.

The analysis of AD scores derived from the prediction model (Figure 4G-I), offers an insightful perspective on the progression of AD when correlating with CDR evaluations. The model shows a significant increase in AD scores when progressing from CDR 0 to 0.5, demonstrating its sensitivity in identifying AD at an early stage. This initial spike in AD score likely reflects the response to the early neuropathological changes characteristic of AD, a phase often associated with the onset of mild cognitive impairment (MCI), a potential precursor to AD. The body’s inherent response to these early disease processes may involve the regulation of specific genes, perhaps as a compensatory mechanism or as a reaction to initial neuronal stress. Conversely, the model shows a gradual decline in AD scores from CDR 0.5 to 3, indicating an inverse trend. As AD advances to more severe stages, this reduction in gene regulation could indicate the disease exacerbation. The genes initially regulated significantly in response to early disease stages may reduce their regulation due to neuronal cell loss and diminished activity in surviving neurons.^43, 44^ Specifically, given that AD is linked with metabolic and bioenergetic alterations in the brain,^45^ and considering the critical role of these genes in energy metabolism, mitochondrial function, and oxidative stress (Figure 2F), it is conceivable that the genes initially enhance their regulatory activity in response to metabolic demands but diminish as neuronal dysfunction intensifies.

In summary, this study provides a comprehensive analysis of mRNA from plasma EVs across 82 AD samples and healthy controls. The application of Next-Generation Sequencing allowed for a detailed gene expression profile, highlighting genetic progression from MCI to AD through differential gene expression and pathway analysis. The functional enrichment and pathway analyses emphasize disruptions in metabolic processes and the role of oxidative stress in neurodegenerative diseases. The study also notes the interconnectedness of neurodegenerative and metabolic disorders, implying that AD pathophysiological processes are closely tied to overall systemic health. This research identified potential biomarkers for AD, and differentiated the biological and molecular functions unique to MCI and AD. The ROC analysis of the AD prediction model demonstrated impressive accuracy, exhibiting a strong correlation with the established CDR evaluation, which indicates the potential of profiling mRNA from plasma EVs as an effective tool for the early detection and continuous monitoring of AD. To further validate the reliability of the identified markers and optimize the prediction model, future studies should focus on analyzing more clinical samples from larger and more diverse cohorts with different ethnicities, genetic backgrounds, and environmental exposures. Conducting longitudinal studies and comparing AD samples with those from other neurodegenerative conditions, such as Parkinson’s disease and Lewy body disease, will better reveal the sensitivity and specificity of this AD prediction tool.

## Materials and Methods

### Clinical specimens

We analyzed a total of 82 plasma specimens, including 25 samples from healthy controls, 13 from individuals with MCI, and 44 from patients diagnosed with AD. Demographics and clinicopathological characteristics of these participants are provided in **Table S1**. These specimens were from three independent patient cohorts: Washington University in St. Louis (Knight ADRC), Indiana University (NCRAD), and PrecisionMed. The study was approved by the institutional review boards of all participating institutions with written informed consent provided by all participants involved in the study.

### EV isolation, library preparation, and RNA sequencing

For isolating EVs, 350 μL of each plasma sample was first diluted with 1×PBS to a final volume of 15 mL. The diluted plasma was then passed through a 0.22 μm syringe filter to remove large particles and aggregates. The isolation of EVs was conducted using the EXODUS platform, which utilized 25 mm-diameter exosome isolation devices (EID) according to the previously published method.^46^ For comparison, the exoEasy Maxi Kit (Qiagen) was employed. Once isolated, the EVs were resuspended in 400 μL of PBS. From this suspension, a 100 μL aliquot of sample was further diluted 4 times for nanoparticle tracking analysis (NTA) using NanoSight (Malvern Panalytical, LM14). The concentration and size distribution of EVs are shown in **Figure S3**.

Another 300 μL of EVs in PBS were then mixed with an equal volume of Dynabeads M-270 Streptavidin beads (ThermoFisher Scientific, 65305), previously conjugated with a biotinylated oligonucleotide that included a PCR handle and a Poly(dT) sequence. These beads were suspended in a lysis buffer containing 0.4% Sarkosyl to facilitate the capture of mRNA from lysed EVs. Following this, solid-phase reverse transcription was performed using Maxima H Minus Reverse Transcriptase (ThermoFisher Scientific, EP0752). The cDNA synthesized on the beads underwent amplification through SMART PCR, using Kapa HiFi Hotstart Readymix (Roche, KK2601), and the resulting DNA was subsequently purified using AMPure XP beads (Beckman Coulter, A63881). The purified DNA was further subject to a second round of PCR and purification. The PCR products then underwent tagmentation using a Nextera XT Library Preparation Kit (Illumina, FC-131-1096). For fragmented mRNA within EVs, NEBNext Small RNA Library Prep Set (New England BioLabs, E7330L) was employed for library preparation according to manufacturer’s instructions. Quality control of the NGS library preparation was accessed via a TapeStation 4200 with High Sensitivity D5000 ScreenTape (Agilent). Finally, RNA sequencing was performed on an Illumina NovaSeq 6000 platform with at least 20M of reads for each library.

### Genome alignment and gene annotation

The sequencing data were first trimmed using Trimmomatic (version 0.4)^47^. Subsequently, these trimmed reads were aligned to the Ensembl reference genome for Homo sapiens (GRCh38.109) using the STAR aligner (version 2.7.10a)^48^. Duplicate reads were identified and eliminated using Picard tools (version 3.0). The raw reads were extracted using the gene transfer format from the Ensembl database corresponding to the Homo sapiens GRCh38.109 genome. This process was facilitated by several R libraries, including GenomicFeatures (version 1.46.5)^49^, GenomicAlignments (version 1.30.0)^49^, and Rsamtools (version 2.10.0)^50^. The reads obtained were then annotated with external gene names and biotypes using the biomaRt package (version 2.50.3)^51, 52^. Low-read counts were filtered out, retaining only those present in at least 20% of individual samples within each of the study groups (*i.e.*, Control, MCI, and AD), with a minimum of 2 read counts per sample. Moreover, any genes that lacked annotations of external gene names were excluded from the analysis.

### Statistical analysis

Differential gene expression analysis was conducted using DESeq2 (version 1.40.2)^53^ supplemented by the sva package (version 3.48.0)^54^ to account for potential hidden batch effect. Criteria for significance were established by requiring a log2 fold chance > |0.5| and adjusted p-value (p-adjusted) < 0.05. The preprocessed data were normalized using DESeq2, followed by a filtering process to retain only those entries where at least 2 samples exhibited normalized values >10 before passing through the sva package and subsequent processes. Following the differential expression analysis, data subjected to variance stabilizing transformation (VST) were retrieved from DESeq2, processed further with the sva and limma packages (version 3.56.2)^55^ for downstream analyses per recommendation outlined in the DESeq2’s vignette.

Functional profiling was then performed using g:Profiler (version e110_eg57_p18_4b54a898)^56^, employing the g:SCS significant threshold with p-adjusted < 0.05. In identifying KEGG AD-related genes from the pool of differentially expressed genes, the analysis utilized statistical domain scope specified as ‘All known genes.’ In contrast, for more focused profiling of these determined AD-related genes, the statistical domain scope was narrowed to ‘Only annotated genes.’

The linear SVM model, implemented in the e1071 package (version 1.7-13)^57^, was employed to develop a diagnostic classifier. This model was paired with a nested cross-validation strategy with inner and outer folds set to 5, facilitated by the caret package (version 6.0-94)^58^, to address class imbalances within the dataset. The classifier’s performance was assessed through ROC analysis, utilizing the pROC package^59^.

For correlation analysis, the biweight midcorrelation from the WGCNA package ^60, 61^ was utilized. Clustering on the heatmap was performed using the ‘complete’ method with Euclidean distance as the measure. Statistical differences in decision values between AD/MCI and Control groups were determined using the Wilcoxon test. Furthermore, the Kruskal-Wallis test was applied to compare decision values across various CDR groups from CDR 0 to 3.

### Visualizations

Volcano plots were produced with GraphBio^62^. Boxplots were constructed using BoxPlotR^63^. The mean ROC curves were generated by RStudio. For the other plots, SRPlot, a web tool for data analysis and visualization, was employed.

## Data Availability

All data produced in the present study are available upon reasonable request to the authors

## Acknowledgements

The authors acknowledge financial support from National Institutes of Health grants (1R41AG076098-01, 1R43AG080878-01, and 2R44GM144009-02). We also thank Knight ADRC of Washington University in St. Louis and their grants [Healthy Aging and Senile Dementia (P01 AG03991), Alzheimer’s Disease Research Center (P30 AG066444), Adult Children Study (P01 AG026276)]. We thank Indiana University for providing samples from the National Centralized Repository for Alzheimer’s Disease and Related Dementias (NCRAD), which receives government support under a cooperative agreement grant (U24 AG21886) awarded by the National Institute on Aging (NIA).

## Supplementary Information AD classification model

The general formulation of the linear SVM classifier can be expressed as:

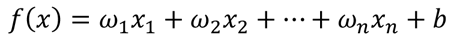

whereas,

- *f(x)* is the decision function of the SVM, which computes a score to predict the class label for a given input vector *x*.
- *ω_1_, ω_2_,…, ω_n_* are the mean weights for each feature (*e.g.*, gene expression levels) in the input vector *x*. These weights represent the average contribution of each gene to the decision boundary, calculated across all iterations.
- *x_1_, x_2_,…, x_n_* are the expression values for a particular gene.
- *b* is the mean bias term, which shifts the decision boundary to enhance the model’s predictive accuracy. It is averaged over 500 iterations.

Specifically, the final model for biomarker panel 1 (Figure 3C), which includes 13 biomarkers, is represented by the following equation:

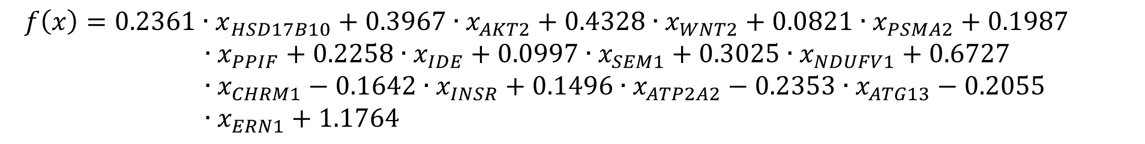

As for biomarker panel 2, which comprises 8 biomarkers (Figure 3D), the equation is as follows:

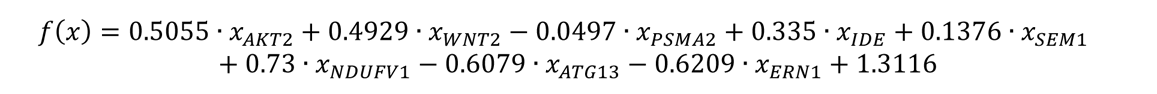

For complementary panel 2, featuring 8 biomarkers (**Figure S2A**), the model is described by the equation:

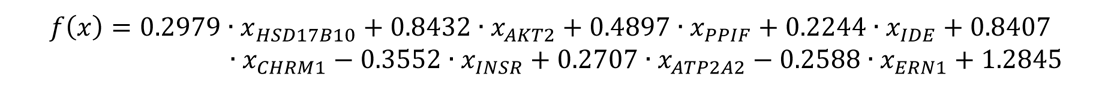

The last panel with 4 biomarkers (Figure 3E), the model is as follows:

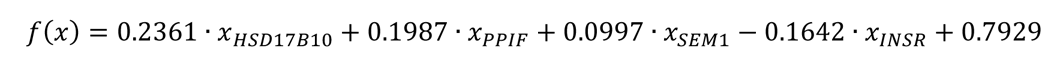

**Table S1.** Information of 82 plasma samples.

| **ID** | **Diagnosis** | **CDR** | **Sex** | **Age** |
| --- | --- | --- | --- | --- |
| 1 | Control | 0 | F | 46-50 |
| 2 | Control | 0 | F | 46-50 |
| 3 | Control | 0 | M | 51-55 |
| 4 | Control | 0 | F | 56-60 |
| 5 | Control | 0 | F | 56-60 |
| 6 | Control | 0 | F | 61-65 |
| 7 | Control | 0 | F | 61-65 |
| 8 | Control | 0 | F | 61-65 |
| 9 | Control | 0 | M | 61-65 |
| 10 | Control | 0 | F | 66-70 |
| 11 | Control | 0 | M | 66-70 |
| 12 | Control | 0 | F | 66-70 |
| 13 | Control | 0 | M | 66-70 |
| 14 | Control | 0 | M | 66-70 |
| 15 | Control | 0 | F | 66-70 |
| 16 | Control | 0 | F | 71-75 |
| 17 | Control | 0 | M | 71-75 |
| 18 | Control | 0 | F | 71-75 |
| 19 | Control | 0 | M | 71-75 |
| 20 | Control | 0 | F | 76-80 |
| 21 | Control | 0 | F | 76-80 |
| 22 | Control | 0 | M | 76-80 |
| 23 | Control | 0 | M | 76-80 |
| 24 | Control | 0 | F | 76-80 |
| 25 | Control | 0 | F | 81-85 |
| 26 | MCI | 0.5 | M | 56-60 |
| 27 | MCI | 0.5 | M | 56-60 |
| 28 | MCI | 0.5 | M | 61-65 |
| 29 | MCI | 0.5 | M | 61-65 |
| 30 | MCI | 0.5 | F | 66-70 |
| 31 | AD | 0.5 | F | 66-70 |
| 32 | AD | 0.5 | F | 66-70 |
| 33 | MCI | 0.5 | F | 66-70 |
| 34 | MCI | 0.5 | M | 71-75 |
| 35 | MCI | 0.5 | M | 71-75 |
| 36 | MCI | 0.5 | M | 71-75 |
| 37 | AD | 0.5 | F | 76-80 |
| 38 | AD | 0.5 | F | 76-80 |
| 39 | AD | 0.5 | F | 76-80 |
| 40 | AD | 0.5 | M | 81-85 |
| 41 | AD | 0.5 | M | 81-85 |
| 42 | AD | 0.5 | F | 86-90 |
| 43 | AD | 1 | M | 56-60 |
| 44 | MCI | 1 | M | 61-65 |
| 45 | AD | 1 | F | 61-65 |
| 46 | AD | 1 | M | 56-60 |
| 47 | MCI | 1 | M | 56-60 |
| 48 | AD | 1 | F | 56-60 |
| 49 | AD | 1 | F | 66-70 |
| 50 | AD | 1 | M | 71-75 |
| 51 | MCI | 1 | M | 71-75 |
| 52 | AD | 1 | F | 71-75 |
| 53 | MCI | 1 | F | 71-75 |
| 54 | AD | 1 | F | 76-80 |
| 55 | AD | 1 | M | 76-80 |
| 56 | AD | 1 | M | 76-80 |
| 57 | AD | 1 | M | 86-90 |
| 58 | AD | 2 | M | 56-60 |
| 59 | AD | 2 | F | 56-60 |
| 60 | AD | 2 | M | 61-65 |
| 61 | AD | 2 | M | 66-70 |
| 62 | AD | 2 | M | 66-70 |
| 63 | AD | 2 | F | 66-70 |
| 64 | AD | 2 | F | 71-75 |
| 65 | AD | 2 | F | 71-75 |
| 66 | AD | 2 | F | 71-75 |
| 67 | AD | 2 | M | 71-75 |
| 68 | AD | 2 | M | 71-75 |
| 69 | AD | 2 | F | 76-80 |
| 70 | AD | 2 | M | 76-80 |
| 71 | AD | 2 | F | 76-80 |
| 72 | AD | 2 | M | 76-80 |
| 73 | AD | 2 | F | 76-80 |
| 74 | AD | 2 | F | 76-80 |
| 75 | AD | 2 | F | 81-85 |
| 76 | AD | 2 | F | 86-90 |
| 77 | AD | 2 | M | 86-90 |
| 78 | AD | 3 | M | 66-70 |
| 79 | AD | 3 | F | 71-75 |
| 80 | AD | 3 | M | 71-75 |
| 81 | AD | 3 | F | 76-80 |
| 82 | AD | 3 | F | 76-80 |

**Figure S1.**
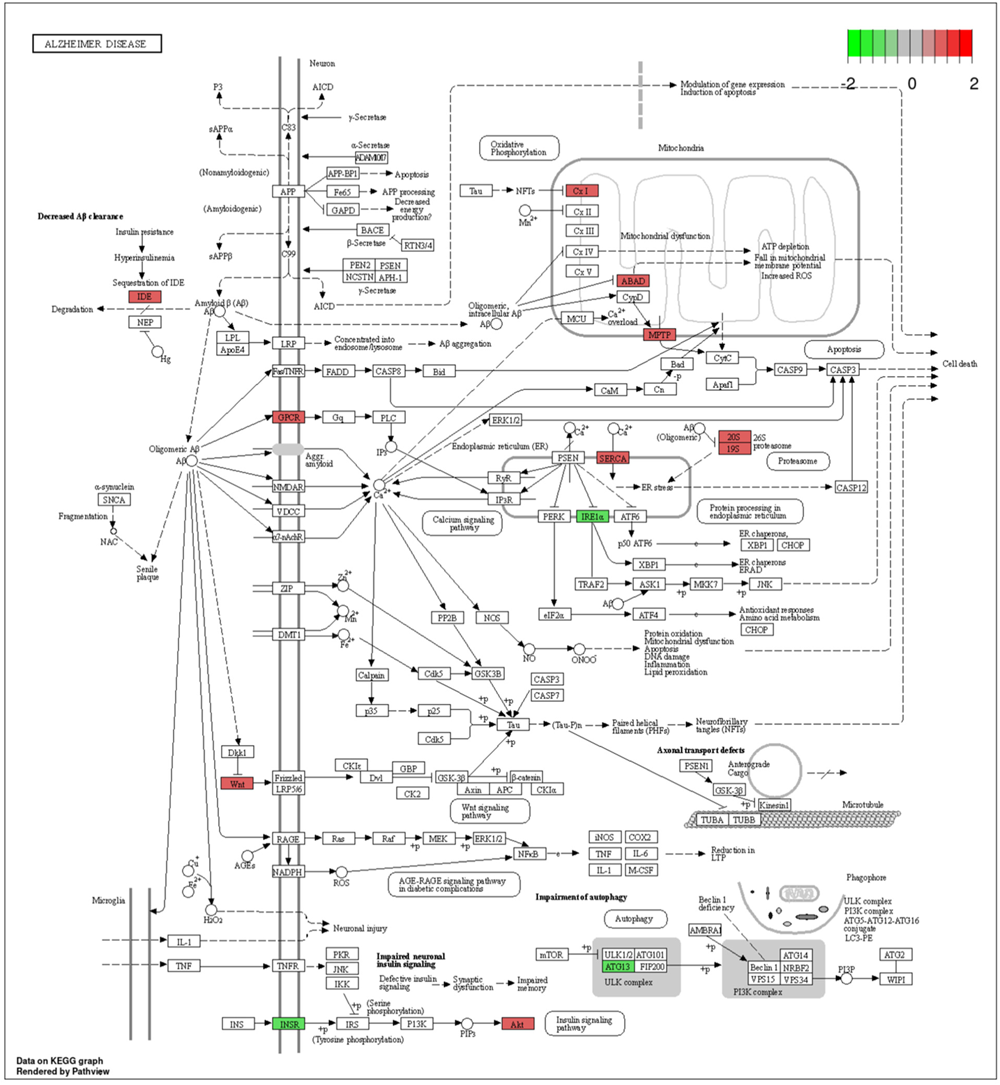
Detailed view into the genes’ involvement of KEGG Alzheimer disease pathway. Red indicates upregulated genes and blue indicates downregulated genes.

**Figure S2.**
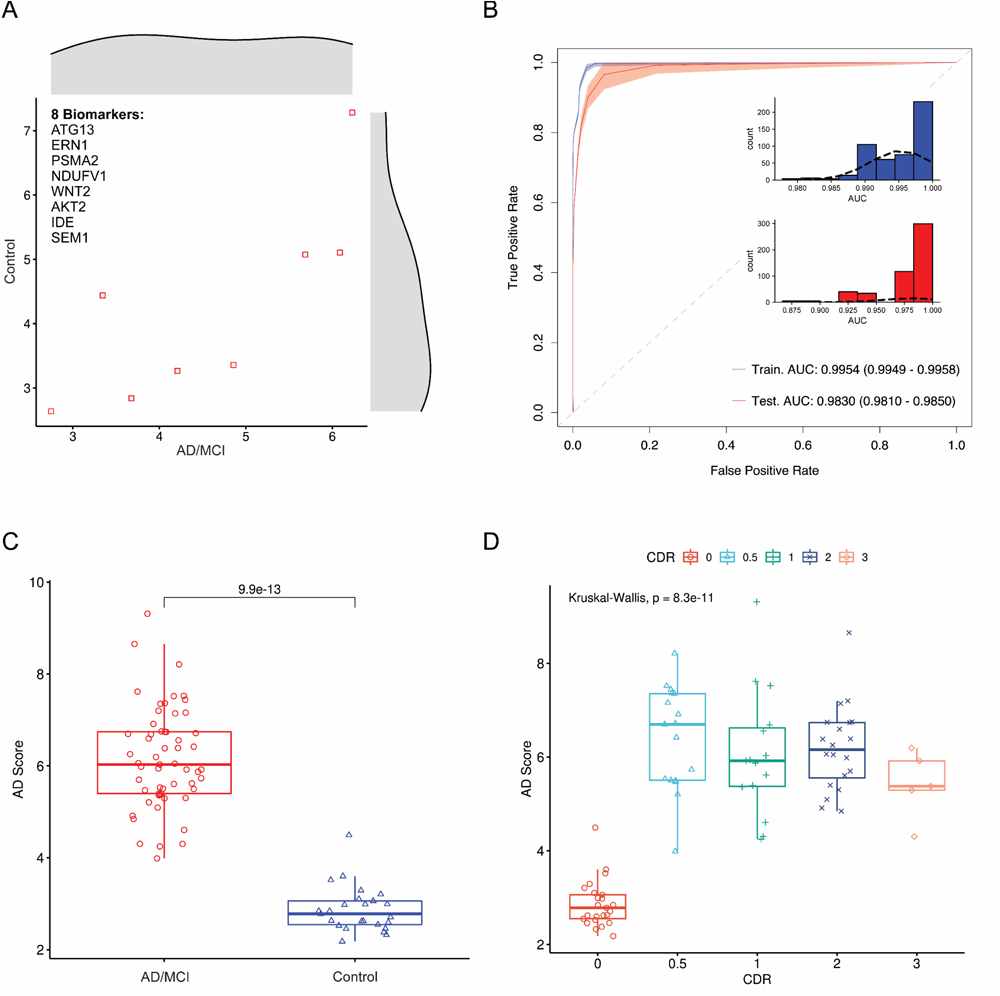
Characterize the performance of an alternative biomarker panel also comprising 8 genetic markers.

**Figure S3.**
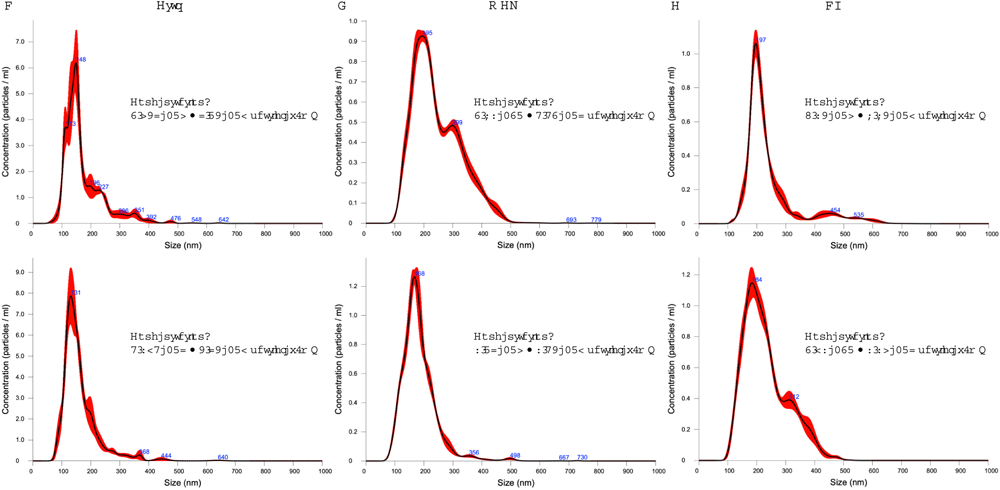
Representative NTA results for (**A**) Control, (**B**) MCI, and (**C**) AD groups.

